# Serological evidence of exposure to filoviruses in bats and humans in Ghana

**DOI:** 10.64898/2026.06.30.26356960

**Authors:** Maya M. Juman, Silke A. Riesle-Sbarbaro, Kofi Amponsah-Mensah, Louise Gibson, Alexandra Mannerings, Yaa Ntiamoa-Baidu, Anthony R. Fooks, Meyir Y. Ziekah, Freya Jephcott, Sylvester Languon, Lewis Drummond, Lianying Yan, Christopher C. Broder, Eric D. Laing, Christian Drosten, Richard D. Suu-Ire, Osbourne Quaye, James L. N. Wood, Andrew A. Cunningham, Olivier Restif

## Abstract

Filoviruses, including the likely bat-borne Ebola virus (EBOV) and Marburg virus (MARV), cause severe hemorrhagic fevers in humans. The 2013-2016 EBOV outbreak caused >11,000 human fatalities in Guinea, Liberia, and Sierra Leone. Nearby countries, including Ghana, have been under sampled for filoviruses relative to West African countries where large outbreaks have occurred. While there have been no reported EBOV disease cases in Ghana, there were two fatal MARV disease cases in 2022, suggesting that at least one filovirus is circulating in the country. In this study, we investigated filovirus circulation in fruit bats and humans in Ghana. We leveraged an extensive serological dataset collected from multiple fruit bat species (*n* = 6,874) and humans (*n* = 1,300) across a decade in Ghana (2010-2020), including both rural and urban regions. We observed evidence of MARV circulation in *Rousettus aegyptiacus* bats, a presumed reservoir, as well as occasional seropositivity in sympatric bat species, suggesting that these other bats are incidental, dead-end hosts. Multivariate analyses suggested that multiple, partially cross-reactive filoviruses are circulating among fruit bats in Ghana. Finally, people who reported spending time in caves and hunting bats had higher serological reactivity against EBOV and MARV relative to individuals with no direct bat exposure, indicating possible undetected filovirus spillover at the bat–human interface in Ghana.

## Introduction

Bats (order Chiroptera) are a geographically widespread and ecologically diverse order of mammals known to host zoonotic pathogens with pandemic potential (Calisher et al. 2006). One family of high-risk zoonotic—and likely bat-borne—RNA viruses is the *Filoviridae* family, which consists of viruses that cause severe hemorrhagic fevers in humans and non-human primates. Members of this family include Ebola virus (EBOV) and Marburg virus (MARV) which circulate in Africa, as well as filoviruses that are non-pathogenic in humans, including Reston virus (RESTV) in Asia (Pourrut et al. 2005). Outbreaks of Ebola virus disease (EVD), a viral hemorrhagic fever caused by EBOV, Bundibugyo virus (BDBV), Sudan virus (SUDV), and Taï Forest virus (TAFV), as well as Marburg virus disease (MVD; caused by MARV), occur sporadically in equatorial African countries. The 2013-2016 EVD outbreak caused over 11,000 human fatalities in Guinea, Liberia, and Sierra Leone (Shultz et al. 2016). Nearby countries, including Ghana, have been under sampled for filoviruses relative to other West African countries with large EVD and MVD outbreaks (Bower and Glynn 2017). While there have been no reported EVD cases in Ghana, models of EVD risk across the African continent have included Ghana in the predicted spillover range (Pigott et al. 2014; Redding et al. 2019). Furthermore, there were two fatal MVD cases in Ghana in 2022, suggesting that at least one filovirus is circulating in the country (WHO 2022).

Predicting and mitigating zoonotic spillover necessitates an understanding of pathogen reservoirs as well as the ecological factors underlying circulation and maintenance in these populations. Many questions remain about the natural history of filoviruses; while RESTV has been identified in Philippine bats (Jayme et al. 2015), Bombali virus (BOMV) has been identified in free-tailed bats (Goldstein et al. 2018; Forbes et al. 2019), and multiple studies have identified the Egyptian fruit bat *(Rousettus aegyptiacus*) as a reservoir host of MARV (Olival and Hayman, 2014), the natural reservoir of EBOV is unknown (Groseth et al. 2007). Serological data provide a starting point for further investigation of this question by offering a valuable proxy for prior infection or exposure (Peel, McKinley et al. 2013). While the specificity and interpretation of serological data can be limited by antibody cross-reactivity, this approach is an important complement to molecular detection of viral nucleic acid material, which is challenging when windows of viremia and active viral shedding are brief (Hayman et al. 2008; 2010; 2012). The detection of filovirus RNA fragments and antibodies reactive against EBOV in some fruit bat species has therefore implicated these bats as possible hosts, though this has yet to be confirmed through virus isolation (Leroy et al. 2005; Pourrut et al. 2007; Hayman et al. 2010).

In EVD outbreak regions in West and Central Africa, the occurrence of several fruit bat species has been shown to increase with human density and footprint, suggesting possible opportunities for bat-human contact and viral spillover (Olivero et al. 2020). One such species is *R. aegyptiacus*. Experimental inoculation studies revealed limited dissemination of EBOV in these bats, suggesting that it is an unlikely reservoir for this virus (Jones et al. 2015; Paweska et al. 2016). However, serology, PCR, virus isolation, and inoculation studies have implicated this species as a reservoir of MARV (Towner et al. 2009). There are particularly high MARV seroprevalence rates in *R. aegyptiacus* cave populations (Pourrut et al. 2009), and experimental infection has confirmed the ability of this species to shed MARV (Amman et al. 2015). Seasonal peaks in MARV seroprevalence in *R. aegyptiacus* living in caves in Uganda correspond to time periods of human MVD outbreaks in this area (Amman et al. 2012).

Another common species of bat that occurs in human areas is the African straw-coloured fruit bat (*Eidolon helvum*), which is broadly distributed across the African continent (Peel, Sargan et al. 2013; Olivero et al. 2020; Cooper-Bohannon et al. 2020). While filovirus RNA has not been detected in this species, multiple studies have found serological reactivity to filovirus antigens in *E. helvum*, suggesting that it might be susceptible to filovirus infection (Ogawa et al. 2015; Lacroix et al. 2021), although experimental work has suggested that these bats’ cells are refractory to EBOV infection (Ng et al. 2015). In one study, an apparently healthy *E. helvum* individual was seropositive for filoviruses and exhibited long-term survival (Hayman et al. 2010). This species’ wide geographic range and highly mobile, nomadic lifestyle create ample opportunities for within- and cross-species viral transmission across at least 42 sub-Saharan African countries (Peel, Sargan et al. 2013; Cooper-Bohannon et al. 2020). Additionally, *E. helvum* is the most heavily hunted bat on the continent and is consumed as wild game in West and Central Africa, creating opportunities for viral spillover into human populations (Kamins et al. 2011; Peel et al. 2017). These bats are often not considered to be a disease risk by communities who hunt and consume them (Kamins et al. 2015). In addition to *E. helvum* and *R. aegyptiacus*, other fruit bat species are common near human areas and remain understudied for filoviruses. For example, bats in the genus *Epomophorus* have shown serological reactivity to filovirus antigens in the Democratic Republic of the Congo, suggesting that these bats warrant further investigation (Lacroix et al. 2021).

In this study, we investigated the extent of filovirus circulation in bats and humans in Ghana. We leveraged a serological dataset collected from multiple fruit bat species and humans across a decade in Ghana (2010-2020). Our human dataset (*n* = 1,300) was collected between 2010 and 2014, spanning 12 urban and rural sites and including samples from bat-hunting populations. Our bat dataset (*n* = 6,874) includes eight longitudinally-sampled roosts (seven wild and one captive) and ten bat species—with a focus on three suspected host species (*R. aegyptiacus*, *E. helvum*, and *Epomophorus gambianus*), sampled from 2012-2020 (Fig. 1). All serum samples were tested for reactivity against EBOV and MARV antigens, and bat samples from 2019-2020 were also tested for reactivity against antigens from eight additional filovirus species: BDBV, RESTV, TAFV, SUDV, BOMV, Lloviu virus (LLOV), Mengla virus (MLAV), and Ravn virus (RAVV). We used these datasets to investigate: 1) filovirus seroprevalence in wild fruit bat populations in Ghana across space, time, and taxonomic boundaries; and 2) the possibility of undetected filovirus spillover into human communities in Ghana. Here, we present analyses of selected subsets of data and publish our complete dataset for future use by other researchers.

**Fig. 1.**
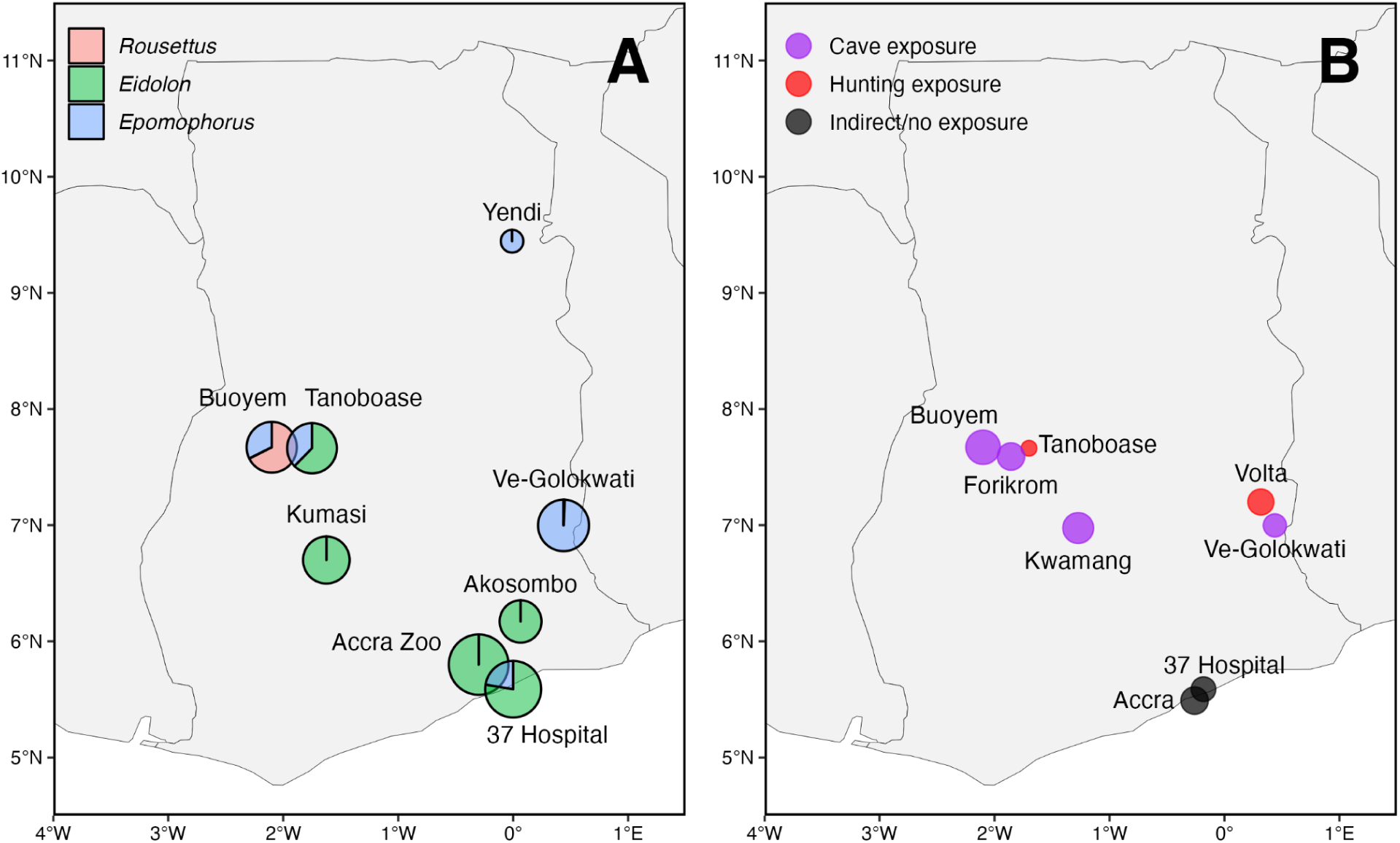
A) Map of bat sampling sites, with point size corresponding to relative sample size and colours indicating the breakdown of the three main species at each site. B) Map of human sampling sites, with point size corresponding to relative sample size and colour indicating exposure. Samples with unknown bat exposure obtained from hospitals were omitted from the map. Smaller villages in the Buoyem area (Akonkonti, Amangoase, and Bonya) are included in the Buoyem site for visual clarity. Coordinates, sampling years, and sample sizes are listed in Supplementary Table 1.

## Results

### Antibody patterns in bats, 2012-2019

We first assessed the overall distributions of corrected EBOV and MARV lnMFI values for wild bats in our dataset (Fig. 2). Using mixture models, we determined conservative species-specific seropositivity cutoffs to use as summary statistics for our dataset (Fig. 2; Supplementary Figs. 1-2). We found that almost 60% of *R. aegyptiacus*, a known MARV host, were seropositive for MARV, and several other species showed some seroreactivity to MARV, including *E. gambianus* (*n* = 1,478, 1.5%), wild *E. helvum* (*n* = 1,050; 9.4%), *Epomops franqueti* (*n* = 42; 36.2%), and *Epomops buettikoferi* (*n* = 21; 52.4%) (Table 1). Some of these incidental positives may be linked to either co-roosting or other contact with *R. aegyptiacus*; for example, *E. gambianus* (the only species with robust sample sizes across sites with and without *R. aegyptiacus* present) had significantly higher lnMFI values for both EBOV and MARV at sites where *R. aegyptiacus* was also sampled, such as Buoyem caves and Ve-Golokwati (Table 2). For example, *E. gambianus* sampled in Buoyem and Ve-Golokwati had mean MARV lnMFI values 0.06 and 0.03 units higher, respectively, than those in Accra where *R. aegyptiacus* is not present (*p* < 0.0001 for both; Table 2; Supplementary Fig. 3).

**Fig. 2.**
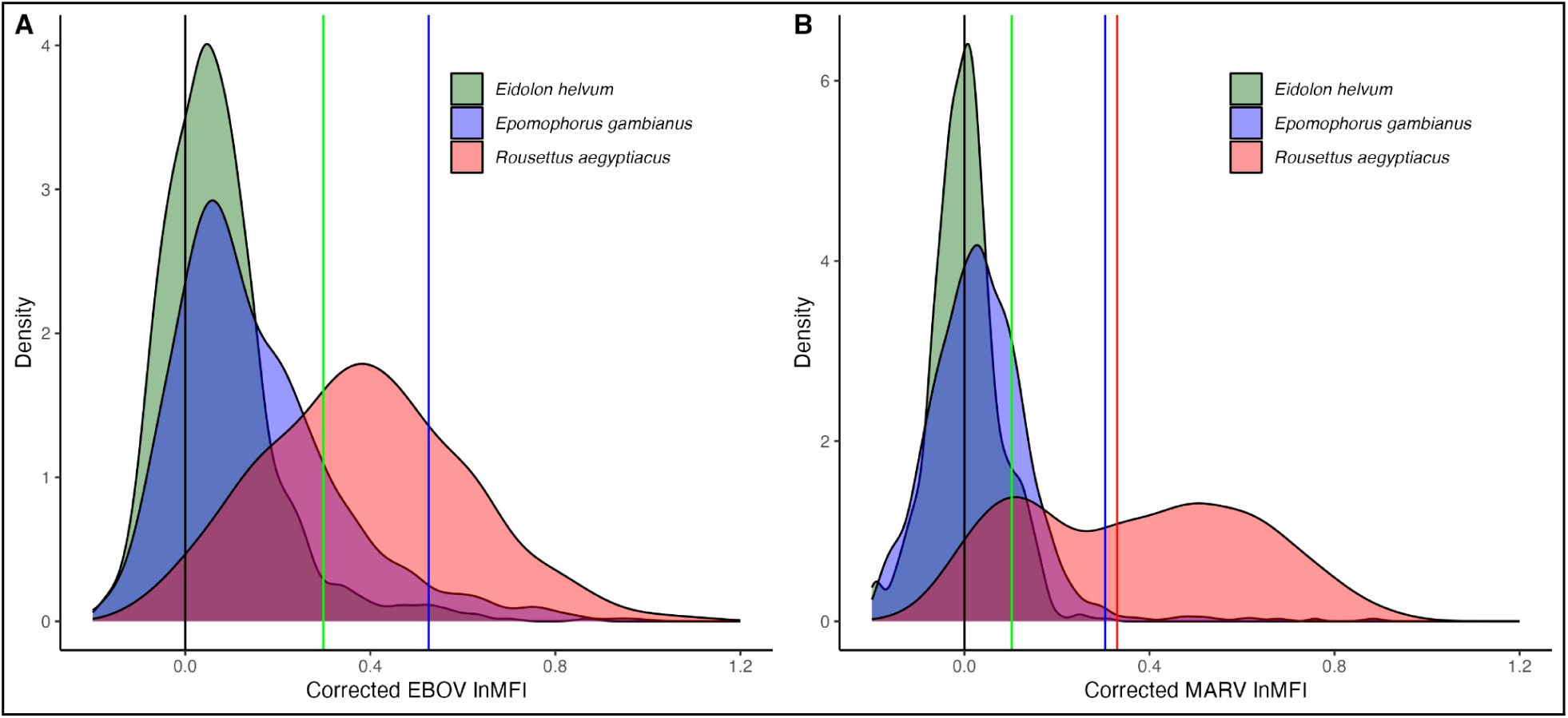
Density plots of corrected lnMFI distributions for the three main species in our dataset (*E. helvum*, *E. gambianus*, and *R. aegyptiacus*) for A) EBOV; and B) MARV. Vertical lines indicate species-specific positivity cutoffs generated by mixture models. The model for EBOV in *R. aegyptiacus* best supported a single cluster, so a cutoff was not determined. Histograms (without density smoothing) are presented in Supplementary Fig. 2.

**Table 1.**
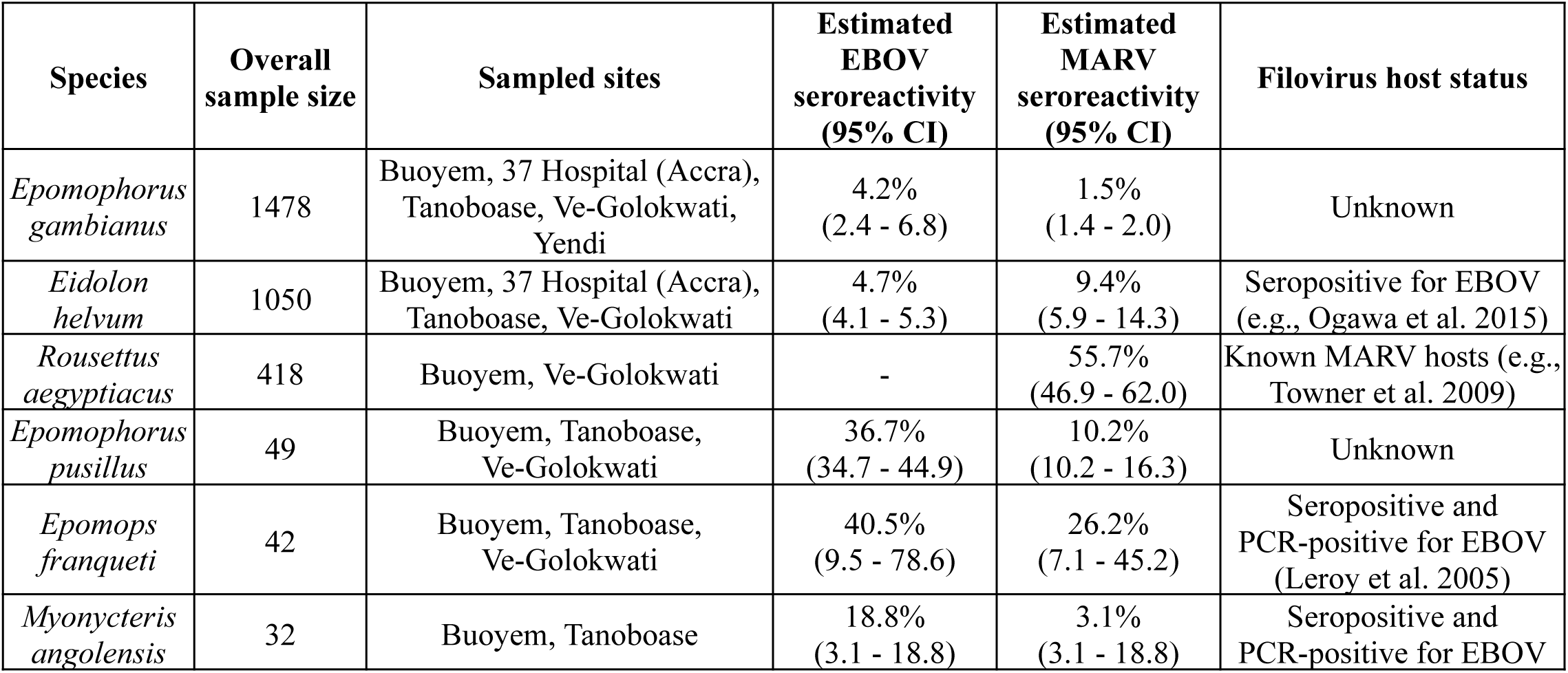
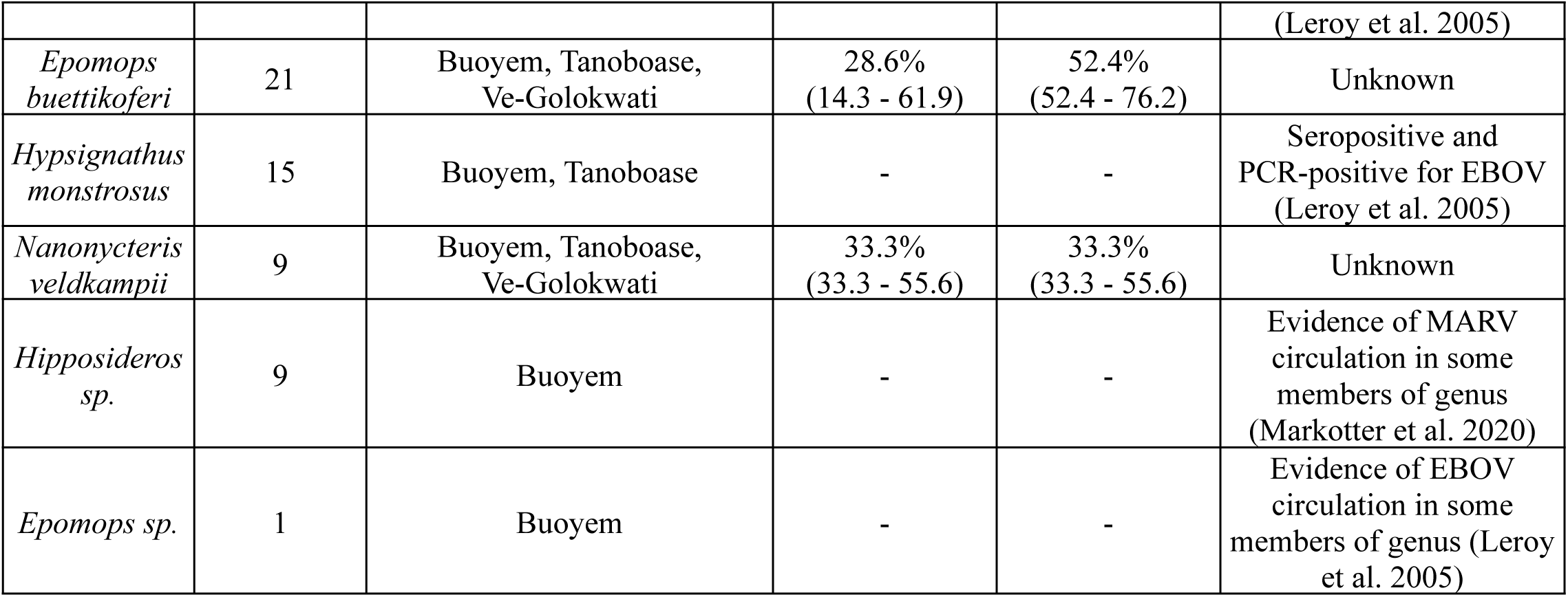
Bat species included in the 2012-2019 dataset and their sample sizes, sites, estimated seroreactivity, and known filovirus associations. For some species (*R. aegyptiacus* EBOV, *H. monstrosus* EBOV and MARV), species-specific seropositivity cutoffs could not be quantified because mixture models identified only one cluster. In the case of *Hipposideros sp.* and *Epomops sp.*, multiple species may have been included in the sample, so cutoffs were not calculated.

| Species | Overall sample size | Sampled sites | Estimated EBOV seroreactivity (95% CI) | Estimated MARV seroreactivity (95% CI) | Filovirus host status |
| --- | --- | --- | --- | --- | --- |
| *Epomophorus gambianus* | 1478 | Buoyem, 37 Hospital (Accra), Tanoboase, Ve-Golokwati, Yendi | 4.2% (2.4 - 6.8) | 1.5% (1.4 - 2.0) | Unknown |
| *Eidolon helvum* | 1050 | Buoyem, 37 Hospital (Accra), Tanoboase, Ve-Golokwati | 4.7% (4.1 - 5.3) | 9.4% (5.9 - 14.3) | Seropositive for EBOV (e.g., Ogawa et al. 2015) |
| *Rousettus aegyptiacus* | 418 | Buoyem, Ve-Golokwati | - | 55.7% (46.9 - 62.0) | Known MARV hosts (e.g., Towner et al. 2009) |
| *Epomophorus pusillus* | 49 | Buoyem, Tanoboase, Ve-Golokwati | 36.7% (34.7 - 44.9) | 10.2% (10.2 - 16.3) | Unknown |
| *Epomops franqueti* | 42 | Buoyem, Tanoboase, Ve-Golokwati | 40.5% (9.5 - 78.6) | 26.2% (7.1 - 45.2) | Seropositive and PCR-positive for EBOV (Leroy et al. 2005) |
| *Myonycteris angolensis* | 32 | Buoyem, Tanoboase | 18.8% (3.1 - 18.8) | 3.1% (3.1 - 18.8) | Seropositive and PCR-positive for EBOV |
|  |  |  |  |  | (Leroy et al. 2005) |
| *Epomops buettikoferi* | 21 | Buoyem, Tanoboase, Ve-Golokwati | 28.6%<br>(14.3 - 61.9) | 52.4%<br>(52.4 - 76.2) | Unknown |
| *Hypsignathus monstrosus* | 15 | Buoyem, Tanoboase | - | - | Seropositive and PCR-positive for EBOV (Leroy et al. 2005) |
| *Nanonycteris veldkampii* | 9 | Buoyem, Tanoboase, Ve-Golokwati | 33.3%<br>(33.3 - 55.6) | 33.3%<br>(33.3 - 55.6) | Unknown |
| *Hipposideros sp.* | 9 | Buoyem | - | - | Evidence of MARV circulation in some members of genus (Markotter et al. 2020) |
| *Epomops sp.* | 1 | Buoyem | - | - | Evidence of EBOV circulation in some members of genus (Leroy et al. 2005) |

**Table 2.** Predictors included in generalised linear models (GLMs) of lnMFI values for EBOV and MARV in wild *E. gambianus*, and their respective estimates, standard errors (*SE*), t statistics (*t* value), and significance levels (*p-*value). The reference site is the 37 Hospital in Accra.

| Antigen | Site | Estimate | SE | *t* value | *p*-value |
| --- | --- | --- | --- | --- | --- |
| EBOV | [Intercept] | 0.13 | 0.01 | 13.15 | <0.0001 |
|  | Buoyem | 0.05 | 0.02 | 2.99 | 0.0029 |
|  | Tanoboase | 0.01 | 0.02 | 0.94 | 0.3486 |
|  | Ve-Golokwati | 0.02 | 0.01 | 1.70 | 0.0901 |
|  | Yendi | 0.04 | 0.04 | 0.91 | 0.3636 |
| MARV | [Intercept] | 0.01 | 0.01 | 1.62 | 0.1060 |
|  | Buoyem | 0.06 | 0.01 | 5.69 | <0.0001 |
|  | Tanoboase | -0.01 | 0.01 | -0.90 | 0.3675 |
|  | Ve-Golokwati | 0.03 | 0.01 | 3.95 | <0.0001 |
|  | Yendi | 0.05 | 0.03 | 1.67 | 0.0954 |

All species for which EBOV cutoffs could be quantified showed some seropositivity, ranging from 4.2% in *E. gambianus* (*n* = 1,478) to 40.5% in *Epomops franqueti* (*n* = 42). Most species showed higher reactivity to EBOV than to MARV, with the exception of *E. helvum* (4.7% EBOV vs. 9.4% MARV) and *Epomops buettikoferi* (28.6% EBOV vs. 52.4% MARV). The wide confidence intervals and small sample sizes for many of the lesser-represented species in our dataset suggest that these cutoffs should be interpreted cautiously and used as summary statistics rather than indicators of true serostatus (Table 1; Supplementary Figs. 1-2). Consequently, we use continuous lnMFI values as response variables in subsequent analyses. There was a statistically significant correlation between overall corrected lnMFI values for EBOV and MARV among wild bats (Pearson correlation = 0.53, *t* = 34.6, *p* < 0.0001; Supplementary Fig. 4).

To assess temporal and demographic patterns in filovirus antibody levels in the three main species studied (*E. helvum*, *E. gambianus*, and *R. aegyptiacus*), we fitted generalised additive mixed models (GAMMs) for both EBOV and MARV. For both antigens, there is a statistically significant effect of time as a smoothing term for all three species, reflecting short-term oscillations (particularly in *R. aegyptiacus*) and an overall increase in lnMFI values beginning around 2017, across all three species (Supplementary Table 2; Fig. 3). The local peaks did not occur at consistent times or seasons across years. In both the EBOV and MARV models, sexually immature bats have corrected lnMFI values 0.02 and 0.03 units lower, respectively, than adult male bats (*p* = 0.01 and < 0.0001, respectively) (Supplementary Table 2). However, female bats (both pregnant and non-pregnant) showed no statistically significant differences from male bats in lnMFI values, and body condition did not have a significant effect on lnMFI values (Supplementary Table 2).

**Fig. 3.**
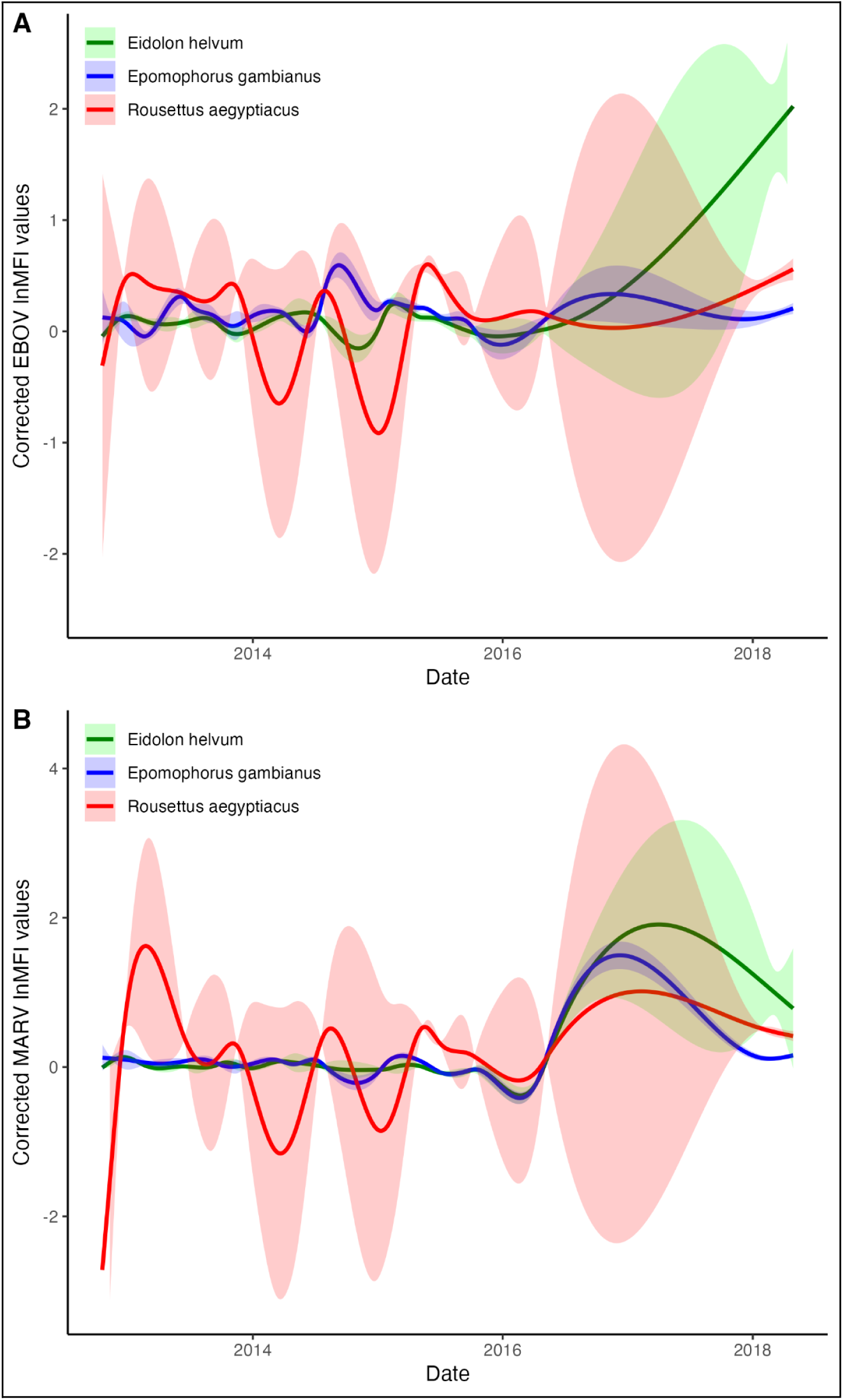
Temporal patterns from GAMMs of A) EBOV and B) MARV corrected lnMFI values for the three main species in our dataset (Supplementary Table 2).

Our 2012-2019 dataset also included repeated sampling of captive *E. helvum* bats in a colony at the Accra Zoo (*n* = 1,641 blood samples from at least 286 resampled bats). Of the bats in this subset, 2.6% (2.3–3.2 95% CI) were seropositive for EBOV and 17.4% (12.1–26.7 95% CI) were seropositive for MARV based on the cutoffs established for wild *E. helvum*, suggesting lower EBOV and higher MARV seroprevalence in the captive colony relative to wild bats. During the sampling period, two female bats displayed marked increases in antibody binding for EBOV (from 0.01 to 0.82 lnMFI and from 0.04 to 0.50 lnMFI) and one male bat displayed a marked increase in antibody binding for MARV (from -0.04 to 0.78 lnMFI) (Supplementary Fig. 5).

### Multivariate analyses of antibodies against ten filovirus antigens in Eidolon helvum, 2019-2020

Corrected lnMFI values for most of the filovirus antigens in our later dataset (exclusively *E. helvum*) were generally less than zero, indicating antigen–antibody binding below the background MFI levels of antibody binding. There was some reactivity across all four sites for EBOV, as well as a cluster of higher BDBV and BOMV lnMFI values in the captive colony and Accra urban roost (Fig. 4). We generated antigen-specific cutoff values using these distributions and found overall seroprevalences of 4.1% (2.9–11.0 95% CI) for BDBV, 20.0% (18.6–23.6 95% CI) for BOMV, and 2.9% (2.5–3.4 95% CI) for EBOV (Supplementary Fig. 6).

**Fig. 4.**
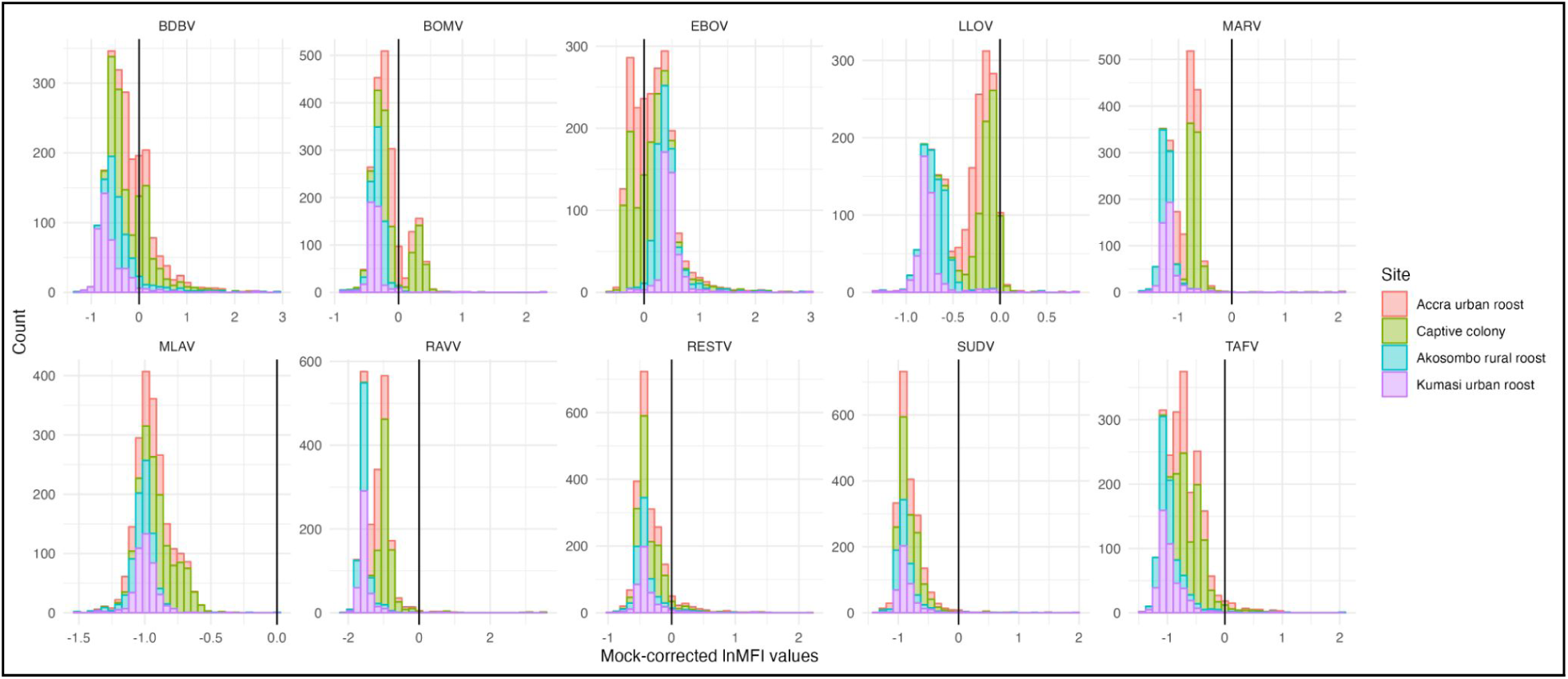
Distribution of corrected lnMFI values for ten filovirus antigens from *Eidolon helvum* bats sampled from 2019-2020.

To assess the degree of cross-reactivity between different filovirus antigens in this multivariate dataset, we calculated Pearson correlation coefficients for all ten antigens. The strongest positive correlations were between MARV and LLOV, MLAV and BOMV, TAFV and BDBV, and RAVV and MARV, all of which were >0.70 (Supplementary Fig. 7). The only negative correlations were between EBOV and LLOV, MARV, RAVV, and MLAV (Supplementary Fig. 7). Principal component analysis of antigen–antibody binding patterns in this expanded ten-antigen panel revealed some spatial structure, where bats in the captive colony and Accra urban roost showed different antigen binding relative to those sampled at Akosombo and Kumasi (Supplementary Fig. 8). These latter bats were more likely to show reactivity to EBOV and none of the other nine antigens, while the captive and wild bats in Accra showed the opposite pattern (Supplementary Fig. 8).

### Human sampling, 2010-2015

There was a statistically significant correlation between EBOV and MARV corrected lnMFI values in the human dataset, suggesting some degree of antigen cross-reactivity in this sample set (Pearson correlation = 0.63, *t* = 29.5, *p* < 0.0001). Humans from communities reporting bat hunting (Volta, Ve-Golokwati, and Tanoboase) or bat interactions in caves (Akonkonti, Amangoase, Buoyem, Bonya, Forikrom, and Kwamang) had slightly, but significantly, higher corrected lnMFI values for both EBOV and MARV than those from populations where there was either no bat contact or indirect contact (Accra) or from febrile hospital patients with unknown bat contact (Noguchi dataset) (Fig. 5). For EBOV, individuals from communities that hunt bats or spend time in caves had respective corrected lnMFI values 0.17 and 0.12 units higher than those with no reported contact (*p* < 0.0001 for both; Table 3). For MARV, individuals from communities that hunt bats or spend time in caves had respective corrected lnMFI values 0.08 units higher than those with no reported contact (*p* < 0.0001 for both; Table 3). Among individuals sampled one year apart from communities that spend time in caves near bats, there was an average increase of 0.02 and 0.07 corrected lnMFI values for EBOV and MARV, respectively, but no individuals surpassed MFI levels of positive controls (Supplementary Fig. 9).

**Fig. 5.**
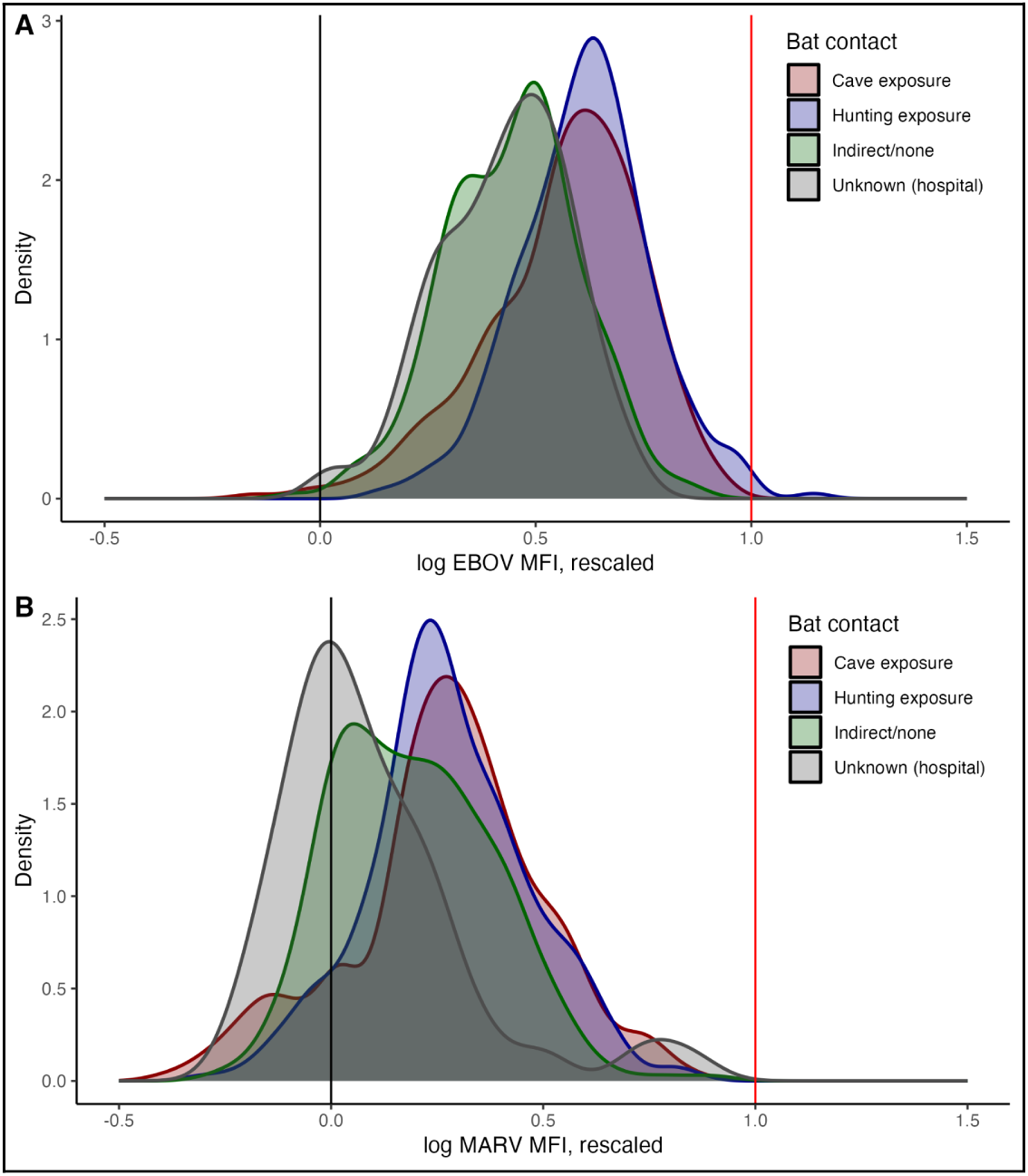
Corrected lnMFI distributions of human samples for A) EBOV; and B) MARV. The red vertical lines indicate the scaled positive control values for EBOV and MARV.

**Table 3.** Exposure types included in generalised linear models (GLMs) of lnMFI values for EBOV and MARV in human populations and their respective estimates, standard errors (*SE*), t statistics (*t* value), and significance levels (*p-*value). The reference exposure level is indirect/none (i.e., volunteers sampled in Accra, with no reported bat contact).

| Antigen | Exposure type | Estimate | *SE* | *t* value | (*p*-value) |
| --- | --- | --- | --- | --- | --- |
| EBOV | [Intercept] | 0.45 | 0.01 | 43.51 | <0.0001 |
|  | Caves | 0.12 | 0.01 | 9.67 | <0.0001 |
|  | Hunting or direct contact | 0.17 | 0.01 | 12.01 | <0.0001 |
|  | Unknown (hospital patients) | -0.02 | 0.02 | -0.82 | 0.4140 |
| MARV | [Intercept] | 0.19 | 0.01 | 15.16 | <0.0001 |
|  | Caves | 0.08 | 0.02 | 5.04 | <0.0001 |
|  | Hunting or direct contact | 0.08 | 0.02 | 4.73 | <0.0001 |
|  | Unknown (hospital patients) | -0.10 | 0.03 | -3.72 | 0.0002 |

## Discussion

Our serological testing results support the occurrence, circulation, and maintenance of MARV in *R. aegyptiacus* cave populations (Towner et al. 2009; Pourrut et al. 2009; Amman et al. 2012; Amman et al. 2015) and constitute the first evidence of *R. aegyptiacus* hosting reactive antibodies to MARV in Ghana, a country which remains understudied for filoviruses relative to other countries in West Africa where larger outbreaks have occurred (Bower and Glynn 2017). The *R. aegyptiacus* population showed antibody reactivity to the MARV and EBOV antigens employed here as early as 2012 (Fig. 3), suggesting that filovirus circulation in West African fruit bats predates the 2013-2016 West Africa EVD outbreak and 2022 MVD cases in Ghana (Shultz et al. 2016; WHO 2022). While the source of these 2022 cases remains unknown, our finding of high MARV seroprevalence in *R. aegyptiacus* bats in Ghana (Fig. 2; Table 1) implies that the MVD patients in 2022 could have been infected within the country (WHO 2022). Similarly, we found evidence of EBOV seroreactivity in at least seven bat species in Ghana; for some species, EBOV seroprevalence was higher than MARV seroprevalence, suggesting that MARV may not be the only filovirus circulating in Ghana (Table 1).

We observed temporal changes in EBOV and MARV antibody binding levels in the three main bat species sampled in our dataset (Fig. 3). Previous work has demonstrated seasonal shifts in MARV seroprevalence in livestock and dogs in Ghana, with an increase during the rainy season (Odoom et al. 2024). Further longitudinal monitoring of bat populations using both serological and molecular approaches will be critical for elucidating whether this seasonal pattern is linked to viral circulation patterns in *R. aegyptiacus*, the presumed MARV reservoir host. Unlike previous studies of other viruses in fruit bats in Ghana (Juman et al. 2024), our study did not reveal any links between reproductive seasonality and filovirus seroprevalence (Supplementary Table 2). However, sexually immature bats had significantly lower EBOV and MARV corrected lnMFI values than adult bats, suggesting that viral prevalence may increase with age (Supplementary Table 2). This age-structured pattern has been identified in other African fruit bat–virus systems, including filovirus circulation in Madagascar (Brook et al. 2019; Juman et al. 2024).

We did not find evidence of widespread filovirus exposure in the other bat species in our dataset, including *Epomophorus gambianus* and *Eidolon helvum*. However, we found some evidence of incidental filovirus occurrence in other species (Table 1). This may suggest that these species show some degree of filovirus susceptibility, serving as occasional, dead-end hosts without providing a long-term reservoir for pathogen maintenance (Odoom et al. 2026). Reactive individuals may have become infected through contact with reservoir species like *R. aegyptiacus*. Indeed, *E. gambianus* bats sampled from sites where *R. aegyptiacus* was present had significantly higher antigen–antibody binding levels than those sampled from sites without *R. aegyptiacus* (Table 2). This suggests that interspecies roost proximity may be linked to cross-species transmission of filoviruses among bats in Ghana, as is the case in other pathogen transmission systems (Hoyt et al. 2018; Simonis and Becker 2026).

Beyond EBOV and MARV, our multivariate analyses suggest that there may be multiple, antigenically-distinct filoviruses circulating in bats in Ghana. This dataset was restricted to *E. helvum*, which had generally low filovirus reactivity; the heavily negative distributions for most of these antigens suggest a lack of filovirus diversity circulating in this species (Fig. 4). However, negative correlations between EBOV and other filovirus antigens (Supplementary Fig. 7) and some spatial separation between the four sampled sites in terms of antigen–antibody binding (Supplementary Fig. 8) suggests the possibility of multiple filoviruses occurring in this species, albeit at low levels. The captive colony and urban roost in Accra show the most similar profile of reactivity, likely because the captive colony was originally populated with bats from the roost in Accra. Bats in the Kumasi and Akosombo roosts showed higher EBOV binding levels (Fig. 4; Supplementary Fig. 8). Together, this suggests that multiple filovirus signals are captured in our sample. This sample includes some reactivity to Bundibugyo and Bombali viruses, which have not been identified in Ghana to date (Fig. 4; Supplementary Fig. 6). Bundibugyo virus has caused human outbreaks in Uganda and the DRC as recently as 2026 (Hulseberg et al. 2021; WHO 2026), while Bombali virus has been identified in wild insectivorous bats in Sierra Leone (Goldstein et al. 2018), Kenya (Forbes et al. 2019; Kareinen et al. 2020), Guinea (Karan et al. 2019), Mozambique (Lebarbenchon et al. 2022), and Tanzania (Düx et al. 2024). Further molecular screening of bats in Ghana will elucidate whether these viruses or related filoviruses are circulating in the sampled populations.

Finally, the results suggest possible undetected filovirus infection in human populations in Ghana with a high degree of bat contact. Individuals from communities exposed to bats through hunting, direct contact (e.g., preparation or consumption of bats), or spending time in caves housing bat roosts had significantly higher corrected lnMFI values for both EBOV and MARV (Fig. 5; Table 3). While the only reported human filovirus disease cases in Ghana occurred in 2022 (WHO 2022), an earlier retrospective serological study revealed 6.3% MARV seroprevalence among humans in Ghana (Steffen et al. 2020). With the exception of febrile hospital patients, samples in our study were drawn from apparently healthy individuals (including those who reported exposure to bats). Taken together, this indicates the possibility of subclinical infection with EBOV, MARV, or antigenically related filoviruses, particularly in populations that interact with bats. Our study suggests that hunting bats or spending time near bats in caves may be risk factors for filovirus infection, a pattern that was similarly noted in a serological study of bushmeat hunters in Guinea (Akoi Boré et al. 2024). Further surveillance of these higher-risk populations is required to definitively confirm this association, particularly in countries with no reported outbreaks or small-scale outbreaks like Ghana, where the extent of filovirus circulation is unknown and the majority of outbreaks may go unreported (Bower and Glynn 2017; Glennon et al. 2019).

In summary, our results suggest that at least one filovirus antigenically similar to MARV is circulating in fruit bat populations in Ghana and may also infect human communities with direct bat or cave exposures. Our study is limited by the use of a potentially cross-reactive assay for antibody detection (Peel et al. 2013). Additionally, the absence of clear multimodal distributions in some species (e.g., EBOV in *R. aegyptiacus*; Fig. S1A) as well as the lack of positive bat controls preclude the estimation of seropositivity cutoff values for all populations. These challenges are inherent to many serological studies, and we interpret our results cautiously in light of the fact that other, related viruses may also be captured by our assay. Future serological surveillance of these populations could benefit from the use of highly multiplexed serology, which has the potential to capture and distinguish between different poorly-characterised viruses (Henson et al. 2023). In conjunction with molecular surveillance of tissue and swab samples from bats and febrile human patients, a broader serological approach may reveal more complex filovirus circulation patterns than those captured in our study.

## Methods

### Bat sampling

We collected 4,784 blood samples from bats across eight sites (seven wild, one captive) in Ghana from 2012 to 2019: 37 Hospital, Accra (*n* = 995), Buoyem DC (*n* = 15), Buoyem MQ (*n* = 114), Buoyem (*n* = 571), Tanoboase (*n* = 657), Ve-Golokwati (*n* = 774), Yendi (*n* = 19), and a captive colony maintained at the Accra Zoo (*n* = 1,639) (Fig. 1A). Sampling was conducted with ethical clearance from the Zoological Society of London Ethics Committee (refs. WLE581, WLE689, WLE715), the Wildlife Division of the Forestry Commission, Ghana, and the Animal and Plant Health Agency, UK. The samples were collected from at least ten bat species, but mostly comprised samples from *E. helvum* (*n* = 2,688), *E. gambianus* (*n* = 1,478), and *R. aegyptiacus* (*n* = 418), with species identification determined visually by morphology (and, in the case of *E. gambianus*, by cytochrome *b* barcoding). An additional 2,090 blood samples were collected exclusively from *E. helvum* bats across four sites (three wild, one captive) in Ghana from February 2019 to October 2020. Sampling was conducted with ethical clearance from the Ghanaian Council for Scientific and Industrial Research (RPN 001/CSIR-IACUC/2018), the Zoological Society of London Ethics Committee (ref. IOZ12), and the United States Army Animal Care and Use Review Office (IACUC protocol # CSIR/IRB/AL/VOL1). The four sites consisted of: 1) Accra urban roost, a seasonally-occupied roost housing up to half a million bats on the grounds of the 37 Hospital (sampled five times, *n* = 496 samples); 2) Kumasi peri-urban roost, a resident peri-urban roost housing up to half a million bats on the grounds of the Kumasi Zoological Garden (sampled four times, *n* = 433 samples); 3) Akosombo rural roost, a roost of unknown population size located in a forested area near the Volta hydroelectric dam (sampled four times, *n* = 386 samples); and 4) a captive colony maintained at the Accra Zoo (sampled five times, *n* = 775 samples including some resampling of the same 179 individuals) (Fig. 1A). Site coordinates, years, and sampled species are listed in Supplementary Table 1.

Wild bats were caught in mist nets while returning to the roost from feeding sites and bats in the captive colony were gathered with landing nets into a large cage. Bats were individually transferred into cotton bags hung from a line until processing. Up to 1 mL of blood (<1% of body weight, less taken from juveniles based on their size) was collected from the cephalic (propatagial) or brachial vein of each bat, and each newly captured bat was banded or microchipped. Age category (adult, subadult, or juvenile), sex, weight (in g), forearm length (in mm), adult female reproductive status (pregnant, lactating, or not reproducing), and identifying microchip number were recorded for each individual. Bats were sorted into one of three age categories, based on morphological characteristics: adult (>24 months), sexually immature (6-24 months), or juvenile (<6 months) (Peel et al. 2016). For bats in the captive colony, year of birth and numerical age were associated with each individual based on their identifying microchip number. Body condition was calculated as weight/forearm length (following Plowright et al. 2008). All wild bats were released at the site of capture immediately after sample collection. Blood samples were processed for serum, and all sera were heat-inactivated at 56 °C for at least 30 minutes before storage.

### Human sampling

We collected 1,300 blood samples from participants across 13 sites in Ghana from 2010 to 2015. Sampling was conducted with ethical clearance from the Zoological Society of London Ethics Committee (WLE581), the IRB of Noguchi Memorial Institute for Medical Research (CPN: 002/10-11), and the Committee for Human Research, Publications and Ethics of Komfo Anokye Teaching Hospital and School of Medical Sciences, Kwame Nkrumah University of Science and Technology (Kumasi). Samples (5 mL) were taken from participants who either had indirect/no bat contact or identified contact with bats, either through hunting or spending time in caves. The sites consisted of: 37 Hospital in Accra (2014; *n* = 120 with indirect or no bat contact), Accra (2010; *n* = 152 with indirect or no bat contact), Akonkonti (2014, *n* = 61 with exposure to caves), Amangoase (2014, *n* = 52 with exposure to caves), Bonya (2015, *n* = 26 with exposure to caves), unspecified Buoyem sites (2014, *n* = 3 with exposure to caves), Tanoboase (2014, *n* = 75 with exposure to bats through hunting), Ve-Golokwati (2014, *n* = 106 with exposure to bats through hunting and direct contact; Ayivor et al. 2017), and Volta (2011, *n* = 135 with exposure to bats through hunting) (Fig. 1B). All blood samples were processed for serum (2 mL), and all sera were heat-inactivated at 56 °C for at least 30 minutes before storage. In addition, 79 samples from acute febrile patients who visited the 37 Hospital in Accra were provided by the Noguchi Memorial Institute for Medical Research and were included in the study. Finally, three sets of matched samples taken one year apart (2011, 2012) from the same individuals were collected from Kwamang (*n* = 119 in 2011, 84 in 2012), Buoyem (*n* = 84 in 2011, 52 in 2012), and Forikrom (*n* = 89 in 2011, 63 in 2012). Individuals from these communities reported spending time in caves near bats (Anti et al. 2015). Site coordinates, years, and sampled species are listed in Supplementary Table 1.

### Serological assays

All serum samples were originally stored in a -20 °C freezer. Bat and human samples were screened for anti-filovirus glycoprotein (GP) antibodies using a multiplex microsphere-based immunoassay (MMIA). Human codon optimised trimeric soluble GP ectodomains were designed as native-like conformations with a GCN_4_-trimerization motif and expressed in stable HEK cell lines using the Freestyle 293 Expression system (Thermo Fisher Scientific, Waltham, MA, USA) (Roe et al. 2025). Affinity purification of S-tag and Twin-Strep-tag GP constructs followed methods previously described (Yan et al. 2023; Cheliout Da Silva 2021). The filovirus GP MMIA was previously validated for human anti-EBOV GP IgG detection (Roe et al. 2025) with correlations to both ELISA and live-EBOV neutralization tests, and it has been adapted for testing blood specimens collected from bat species (Laing et al. 2018; Redekal et al. 2025). Briefly, serum samples were diluted (1:50) and incubated with antigen-coupled microspheres (for 30 minutes on a shaking platform at medium speed), and biotinylated-Protein A (Thermo Scientific, Rockford, IL, USA) and biotinylated-anti-human IgG (Thermo Scientific, Rockford, IL, USA, Pierce Goat Anti-Human IgA + IgG + IgM, (H+L), biotin conjugated) were used as secondary conjugates for bat and human samples, respectively. After washing, antigen-antibody complexes were incubated with streptavidin-phycoerythin followed by a final wash step (1X PBS-T). Subsequently, 50 μL solution was added to each well of the 96-microtiter plate, and resuspended antigen-antibody complexes were measured on Luminex xMAP multiplexing systems and reported in median fluorescence intensity (MFI) units. All samples were screened for EBOV and MARV antibodies. Antigen-antibody binding levels (EBOV and MARV) for 2012-2019 samples were measured via a Luminex 100/200 instrument (LiquiChip, QIAGEN, Germany). Bat samples collected between 2019 and 2020 (*n* = 2,090) were additionally screened against a panel of eight more filovirus antigens (BDBV, RESTV, TAFV, SUDV, LLOV, MLAV, BOMV, and RAVV) along with a mock bead with no viral antigen, using a Luminex FLEXMAP 3D instrument and biotinylated-Protein A and biotinylated-Protein G (Thermo Scientific, Rockford, IL, USA) as secondary conjugates. The 2012-2019 and 2019-2020 datasets were not combined in further analysis due to the different Luminex multiplexing machines used to process them and the different sets of antigens included.

All MFI values were log-transformed (using natural logarithm) prior to analysis to reduce over-dispersion. For the samples processed without a mock bead (i.e., all human and bat samples collected until 2019), each lnMFI value was normalised between 0 and 1 by calibrating with a blank well (transformed to 0) and a positive control (transformed to 1). For EBOV, Monkey 09035 EBOV+, or if unavailable, a monoclonal antibody (mAb 5D2 or mAb 5E6 at 1:50 dilution), was used as a positive control. For MARV, Monkey 90B025 MARV+ or Monkey R0559 MARV+, or if unavailable, a monoclonal antibody (mAb 2B8 or mAb 3B6 at 1:50 dilution), was used as a positive control. This correction produces lnMFI distributions that are mostly between 0 and 1, with some negative values (indicating that antigen-specific antibody binding is lower than binding in blank wells) and some values greater than 1 (indicating that antigen-specific antibody binding is higher than that of a positive control sample). For those with a mock bead (i.e., bat samples from 2019 to 2020), each lnMFI value was corrected by subtracting the background lnMFI value for each sample’s baseline antibody binding to mock beads. This correction produces lnMFI distributions that include some negative values indicating that, in these cases, antigen-specific antibody binding is lower than the background binding levels.

### Statistical analyses

All analyses and visualizations were conducted in R (version 4.2.1; R Core Team, 2022) using the *ggplot2* package (Wickham 2016). Correlations between different antigens were assessed with Pearson correlation coefficients and visualised using the *corrplot* package (Wei and Simko 2021). To quantify seroprevalence to EBOV and MARV, we used the package *mclust* to apply Gaussian finite mixture models to corrected lnMFI distributions of each species and separate individuals into different serostatus categories (Scrucca et al. 2023). The best-fitting model was selected for each distribution using BIC. When two clusters were identified, these were treated as “seronegative” and “seropositive”; when the best-fitting model included three clusters, these were considered to be “background MFI,” “seronegative,” and “seropositive.” Conservative cutoff values were calculated as the mean of the “seronegative” distribution plus three times its standard deviation. When only one cluster was identified, no cutoff value was calculated. We performed nonparametric bootstraps (with 999 replications) of the mixture models to generate 95% confidence intervals for these cutoff values. We then applied the resulting species-specific cutoffs to corrected lnMFI values to quantify the approximate number of “positive” individuals. We emphasise that these cutoffs were used for analytical convenience of separating bats with relatively low or high MFI values and estimating seroprevalence, not as evidence of exposure to specific antigens or viruses or for determining immune status for a specific pathogen.

Subsequent analyses focused on a subset of the larger sample set, comprising only *R. aegyptiacus*, *E. helvum*, and *E. gambianus* (*n* = 4,584). We fitted generalised linear models (GLMs) of *E. gambianus* (*n* = 1,478) with site as a predictor, to determine whether bats at sites with *R. aegyptiacus* present (e.g., Buoyem caves, Ve-Golokwati) were more likely to have higher lnMFI values for EBOV or MARV than those at sites without *R. aegyptiacus* (this could not be repeated for *E. helvum*, as very small numbers of *E. helvum* were sampled where the two species are sympatric). We then fitted generalised additive mixed models (GAMMs) using the *mgcv* package (Wood 2017) to explore the effect of demographic factors and variation in EBOV and MARV antibody binding over time in the three main species in our dataset. We used penalised thin-plate regression splines to fit sampling date as a nonlinear smoothing term for each species, with demographic status (adult male, adult female, pregnant/lactating female, sexually immature, female with unknown reproductive status) and body condition as fixed effects and corrected EBOV and MARV lnMFI values as response variables. For *E. helvum* bats in the captive colony, we plotted the individual trajectories of resampled male and female bats of all ages (*n* = 1,249) to visualise longitudinal changes in filovirus antibody binding in a closed community.

For the ten-antigen Luminex panel (2019-2020 *Eidolon helvum* samples; *n* = 2,090), we used Pearson’s correlation coefficient and principal component analysis to examine multivariate patterns in antigen–antibody binding across the four sites. For antigens with high MFI values, we used the mixture modeling approach described above to generate antigen-specific cutoff values and quantify seropositivity. For the human samples, we visualised differences in corrected lnMFI values between populations with different types of bat exposure using a density plot. We then fitted GLMs of corrected EBOV and MARV lnMFI values with exposure type as a predictor to determine whether individuals who hunted bats or spent time in caves had higher lnMFI values than those with indirect or unknown exposure to bats.

In all statistical models, the threshold for statistical significance was adjusted with a Bonferroni correction to account for testing multiple hypotheses on the same dataset with correlated lnMFI response variables. When two models were fitted to the 2012-2019 bat data or human data (two filovirus antigens), the cutoff for significance was set at *p* = 0.025.

## Data and code availability

All raw data are provided in the supplement, including bat data from 2012-2019 (Supplementary Data S1), bat data from 2019-2020 (Supplementary Data S2), and all human data (Supplementary Data S3). Code for processing and analyzing the raw data is housed in a publicly available GitHub repository, along with the data files (https://github.com/mayajuman/filoviruses/).

## Acknowledgements

We thank the communities who participated in this research and generously donated their time and blood samples—this study would not have been possible without them. This research was developed with funding from the Defense Advanced Research Projects Agency (DARPA) administered through Cooperative Agreement D18AC00031-PREEMPT, as well as with support from the Dynamic Drivers of Disease in Africa Consortium (DDDAC; NERC project no. NE-J001570-1) and Deutsche Forschungsgemeinschaft within the Africa Infectious Diseases program. M.M.J. and A.M. were supported by Gates Cambridge Scholarships enabled by grant OPP1144 from the Bill & Melinda Gates Foundation. J.L.N.W. and O.R. are funded by The Alborada Trust. We are grateful for gifts of positive control sera and monoclonal antibodies from Dr. Thomas Geibert (Galveston National Laboratory, The University of Texas Medical Branch) and Dr. Gary Kobinger (Special Pathogens Program, National Microbiology Laboratory, Public Health Agency of Canada), respectively. We thank the Forestry Commission of Ghana for permitting the collection of the samples and the bat keepers at the Accra Zoo for their vital assistance, as well as Sophia Chrysostomou for assistance with lab records. The opinions and assertions expressed herein are those of the authors and do not reflect the official policy or position of the Uniformed Services University, the Henry M. Jackson Foundation for the Advancement of Military Medicine, Inc., or any other agency of the United States government.

