## Supplementary Information for "Serological evidence of exposure to filoviruses in bats and humans in Ghana"

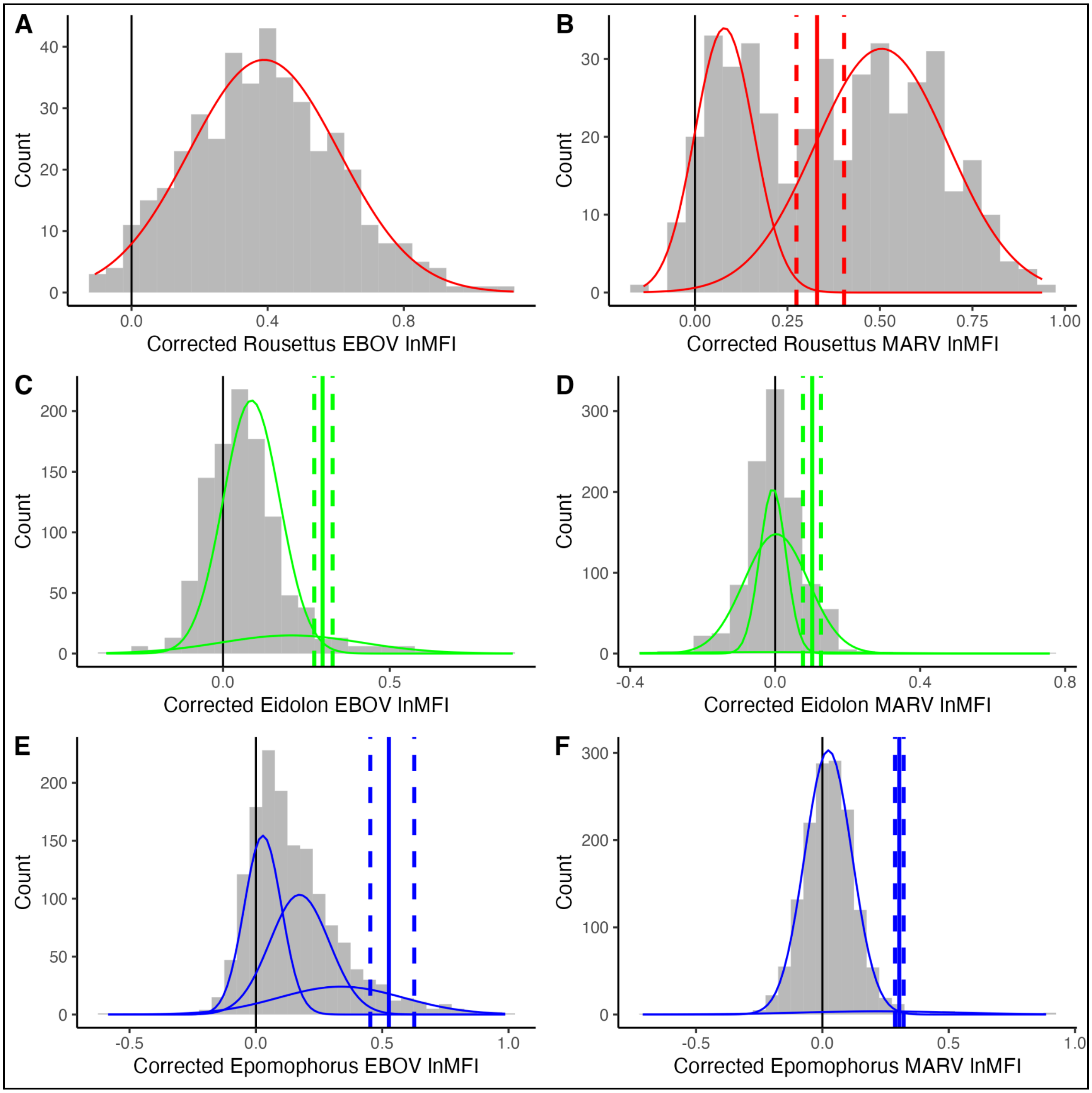


**Supplementary Fig. 1.** Histograms of lnMFI distributions for: A) *R. aegyptiacus* EBOV; B) *R. aegyptiacus* MARV; C) *E. helvum* EBOV; D) *E. helvum* MARV; E) *E. gambianus* EBOV; and F) *E. gambianus* MARV. Coloured curves indicate clusters from the best-fitting mixture model, while coloured vertical lines indicate seropositivity cutoff values (dashed lines indicate 95% confidence intervals, obtained through bootstrapping). For *R. aegyptiacus*, the best fitting EBOV model included only one cluster, so no cutoff value was calculated.

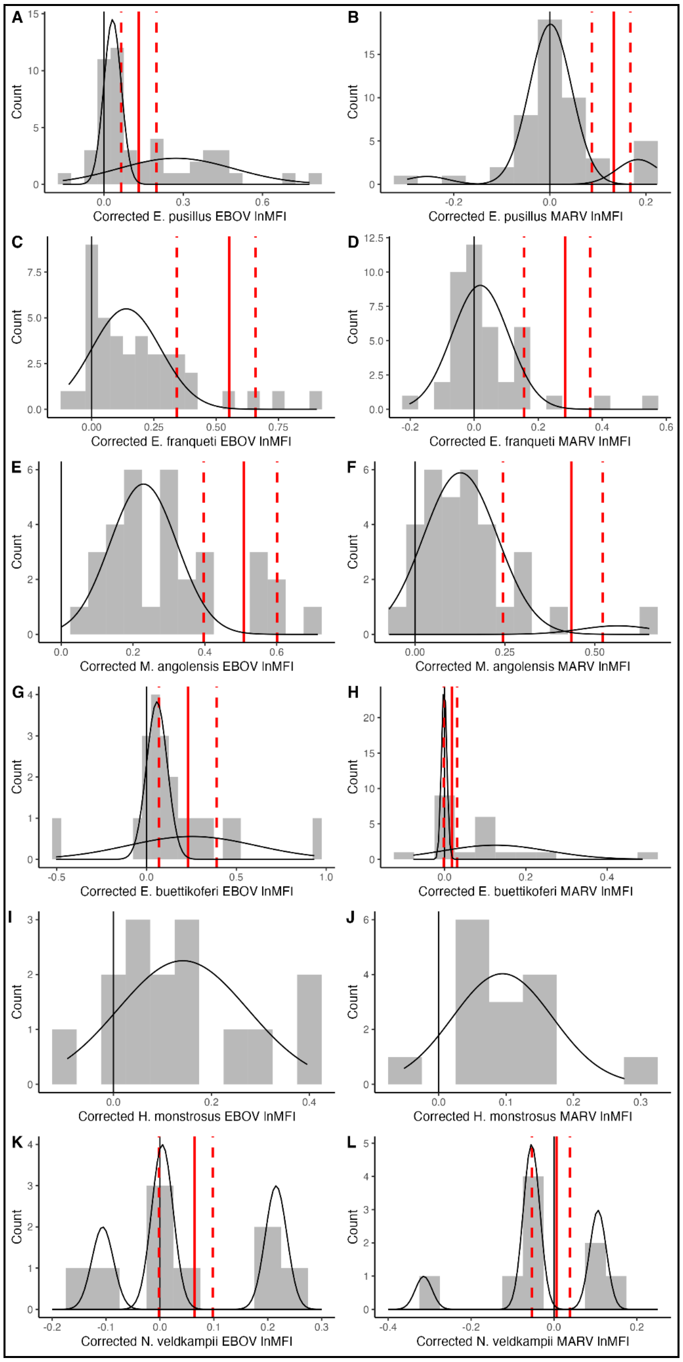


**Supplementary Fig. 2.** Histograms of lnMFI distributions for other species in the dataset: A) *E. pusillus* EBOV; B) *E. pusillus* MARV; C) *E. franqueti* EBOV; D) *E. franqueti* MARV; E) *M. angolensis* EBOV; F) *M. angolensis* MARV; G) *E. buettikoferi* EBOV; H) *E. buettikoferi* MARV; I) *H. monstrosus* EBOV; J) *H. monstrosus* MARV; K) *N. veldkampii* EBOV; and L) *N. veldkampii* MARV. Black curves indicate clusters from the best-fitting mixture model, while red vertical lines indicate seropositivity cutoff values (dashed lines indicate 95% confidence intervals, obtained through bootstrapping). For some species, the best fitting models included only one cluster, so no cutoff values were calculated.

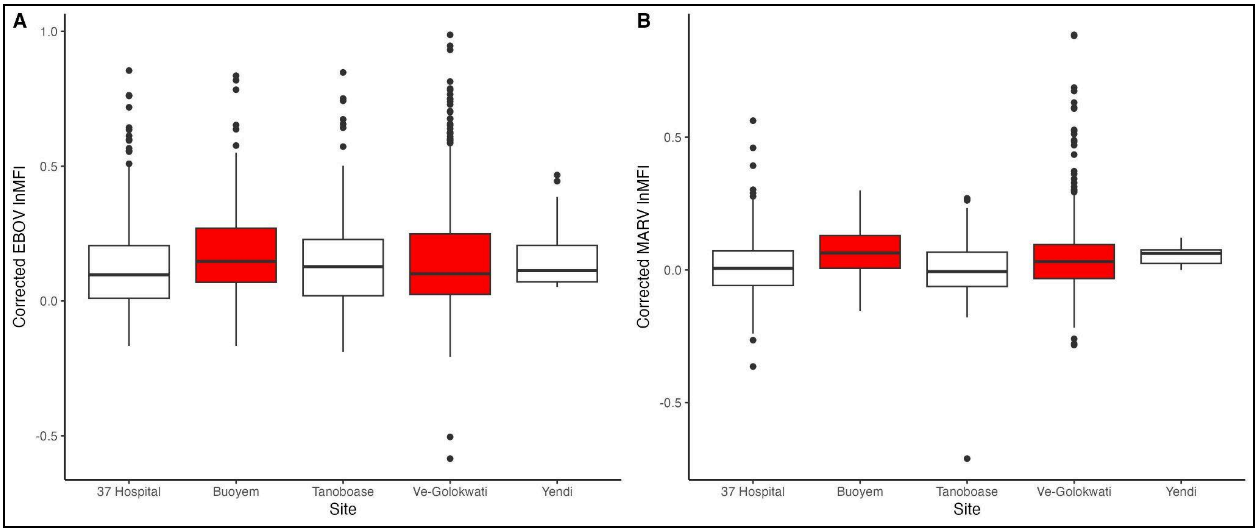


**Supplementary Fig. 3.** Boxplots of *E. gambianus* corrected A) EBOV and B) MARV lnMFI values by collection site. Red boxes indicate sites where *R. aegyptiacus* is also present.

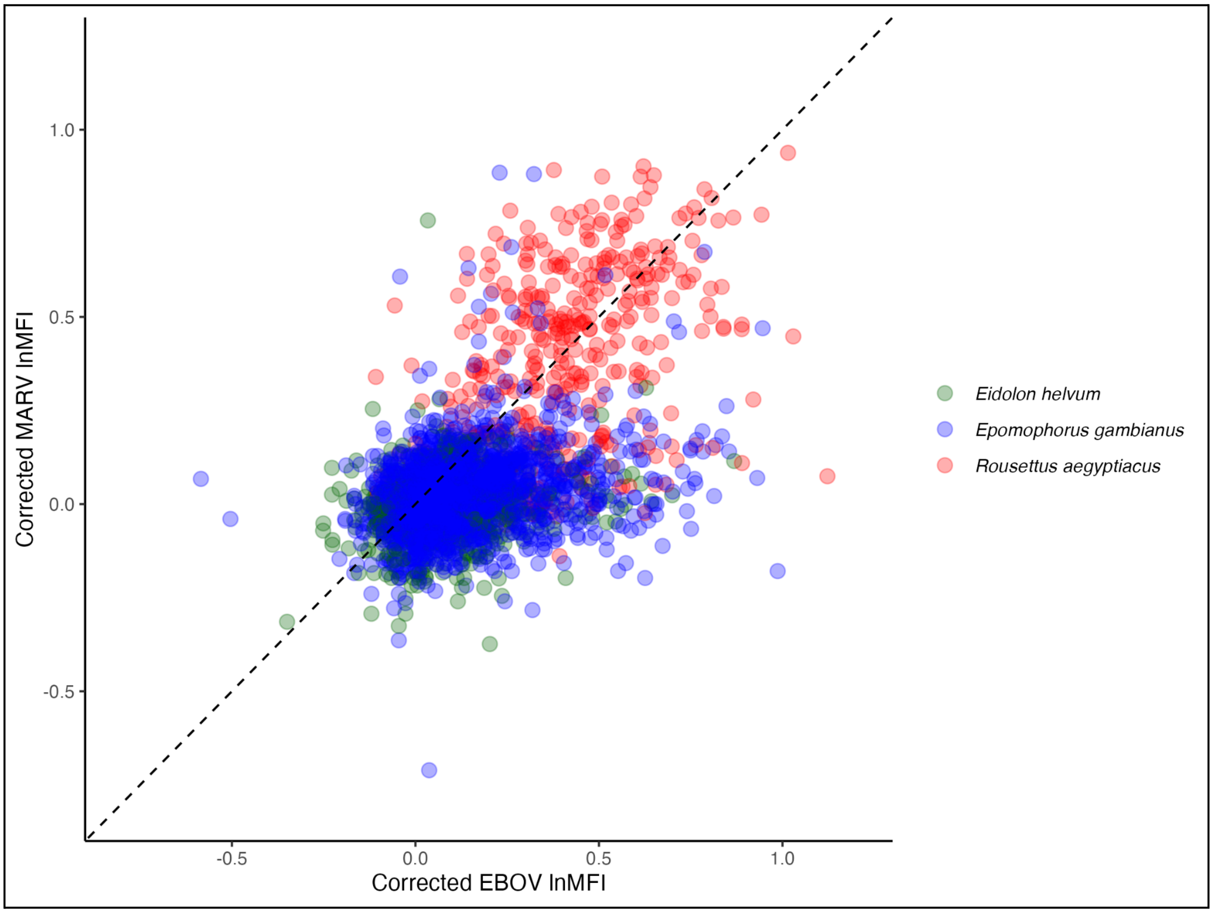


**Supplementary Fig. 4.** Scatterplot of corrected lnMFI values for EBOV and MARV from wild bats sampled from 2012-2019, colour coded to indicate the three main species in our dataset.

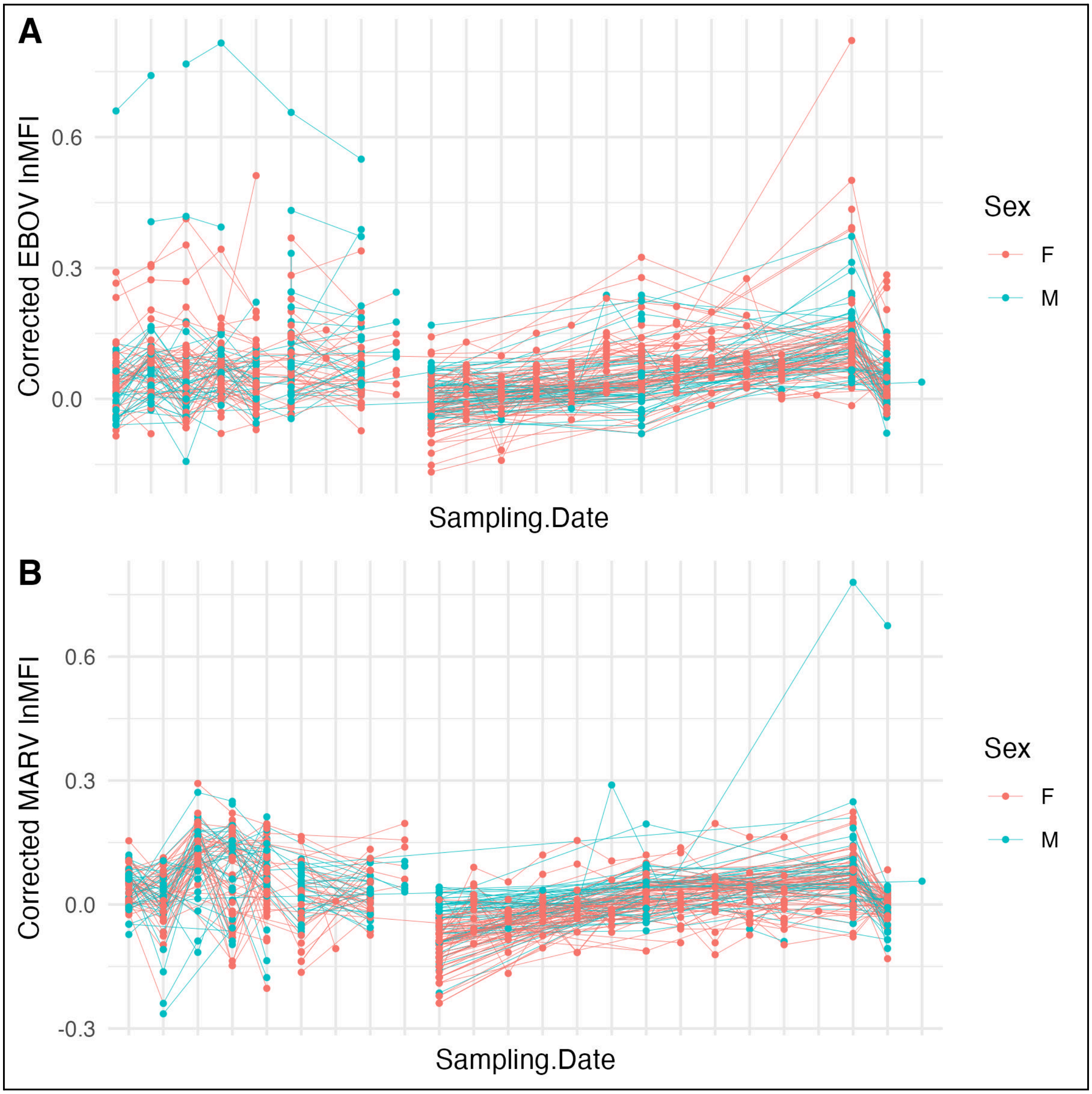


**Supplementary Fig. 5.** Corrected lnMFI values for A) EBOV and B) MARV in the captive colony of *E. helvum* between 2012 and 2019, colour coded by sex. Connected points indicate individuals that were repeatedly sampled.

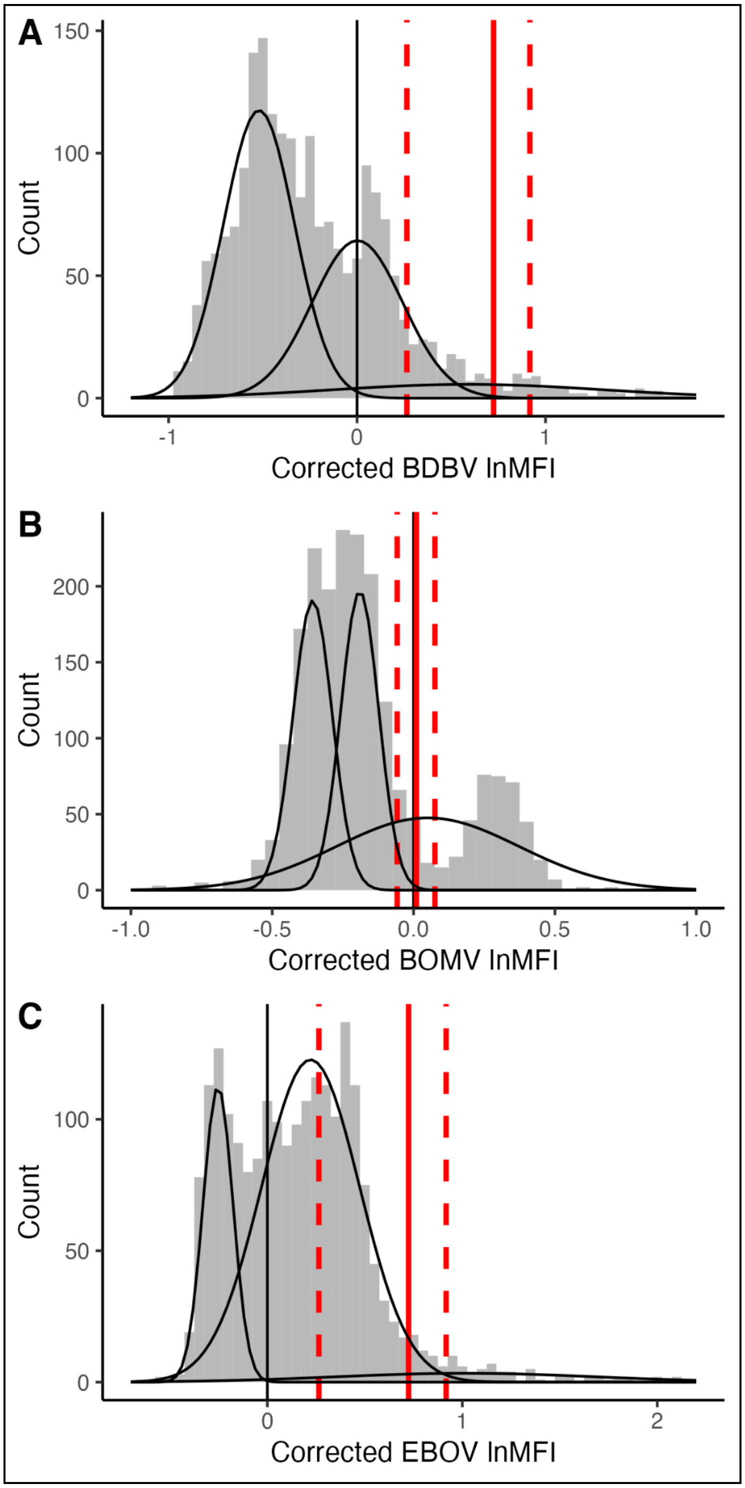


**Supplementary Fig. 6.** Histograms of lnMFI distributions for: A) Bundibugyo virus (BDBV); B) Bombali virus (BOMV); and C) Ebola virus (EBOV) in the 2019-2020 *E. helvum* dataset. Black curves indicate clusters from the best-fitting mixture model, while coloured vertical lines indicate seropositivity cutoff values (dashed lines indicate 95% confidence intervals, obtained through bootstrapping).

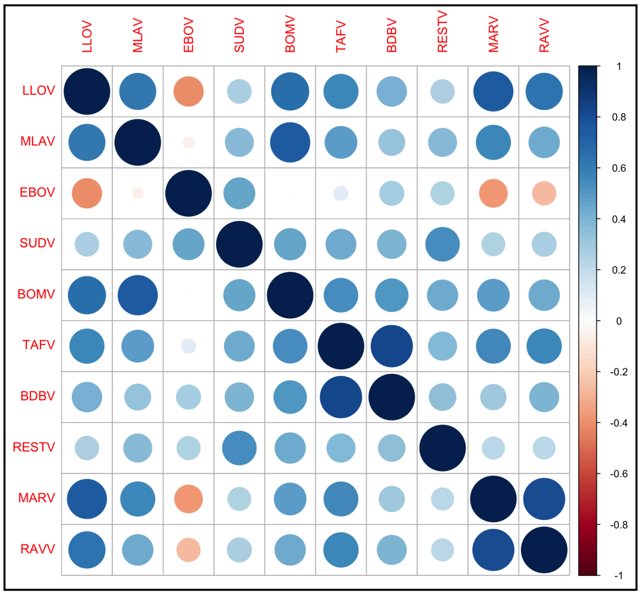


**Supplementary Fig. 7.** Correlation plot showing the magnitude of Pearson correlation coefficients for the ten filovirus antigens included in the Luminex panel for *E. helvum* bats sampled between 2019 and 2020.

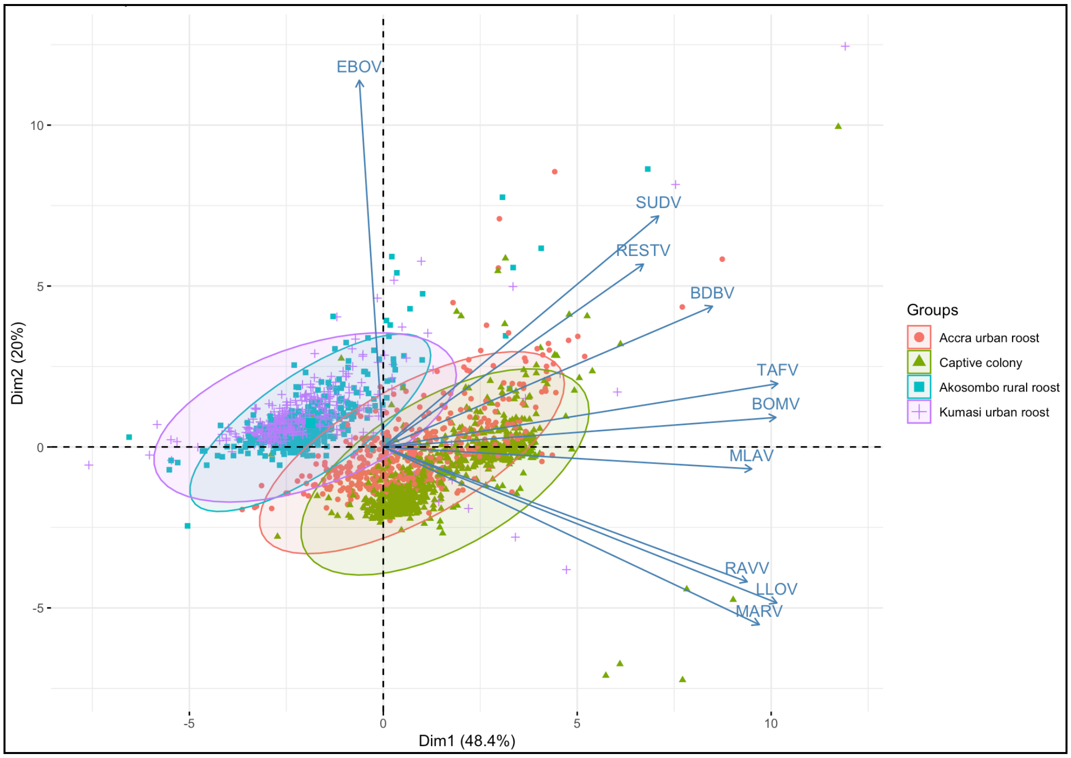


**Supplementary Fig. 8.** Principal component analysis showing patterns in multivariate antigen binding across the four sites where *E. helvum* was sampled from 2019-2020. Ten filovirus antigens were included in the Luminex panel.

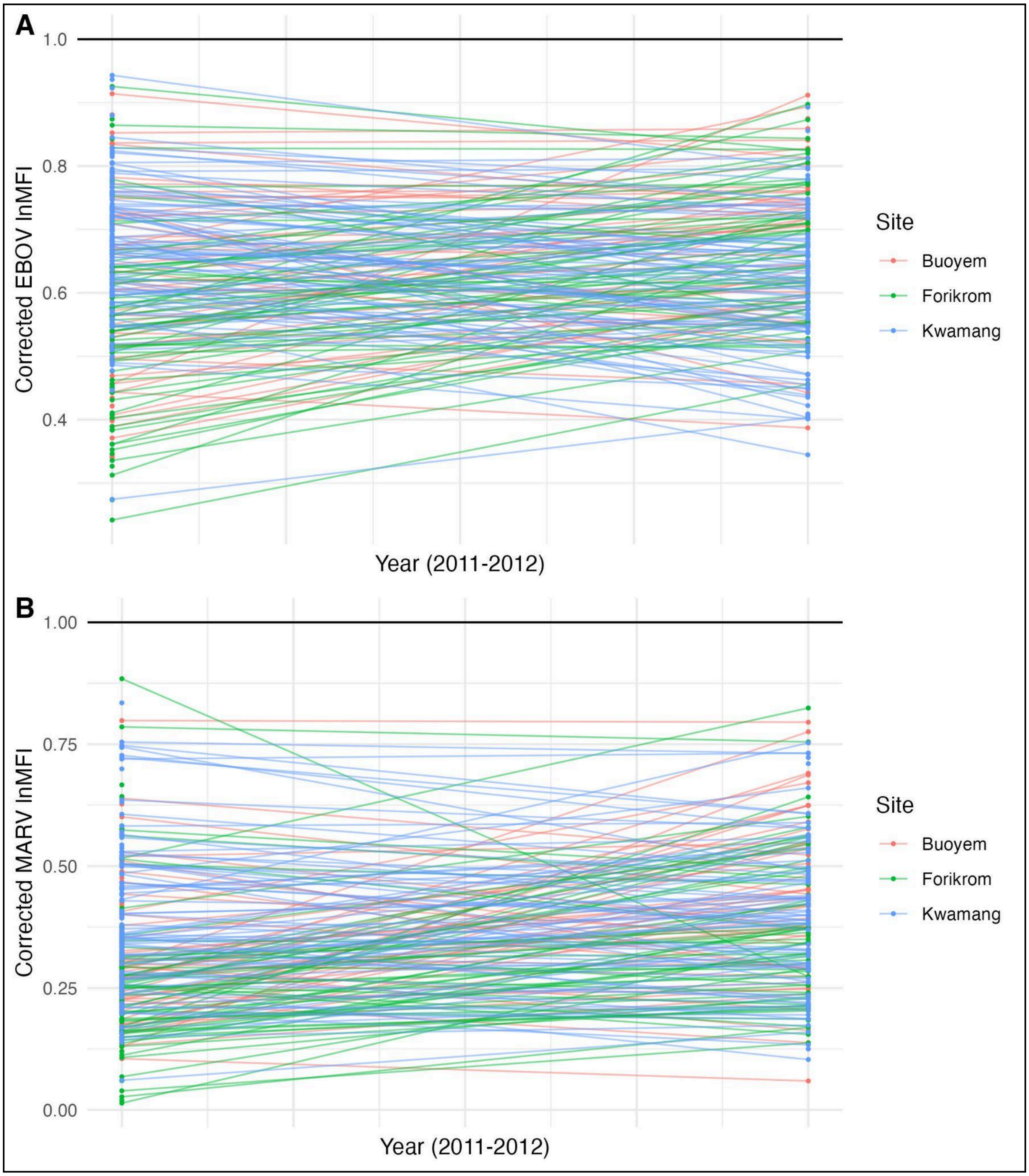


**Supplementary Fig. 9.** Corrected lnMFI values for A) EBOV and B) MARV from human individuals sampled in Buoyem, Forikirom, and Kwamang in 2011 and again in 2012. All individuals were from communities reporting contact with bats in caves. Black horizontal lines indicate the scaled positive control values for EBOV and MARV.

**Supplementary Table 1.** Characteristics and locations of sampling sites included in the study.

| Site | Coordinates | Sampling years | Bat species or human exposure type | Total *n* |
|---|---|---|---|---|
| Bat samples (tested against EBOV, MARV) | | | | |
| 37 Hospital, Accra | 5.5882, -0.18239 | 2012, 2013, 2014, 2015, 2016 | *Eidolon helvum*, *Epomophorus gambianus* | 995 |
| Buoyem (MQ, DC, and unspecified caves) | 7.666667, -1.950000 | 2012, 2013, 2014, 2015, 2016, 2018 | *Eidolon helvum*, *Epomophorus gambianus*, *Epomops franqueti*, *Epomops buettikoferi*, *Epomops sp.*, *Hypsignathus monstrosus*, *Hipposideros sp.*, *Epomophorus pusillus*, *Myonycteris angolensis*, *Nanonycteris veldkampii*, *Rousettus aegyptiacus* | 700 |
| Tanoboase | 7.665417, -1.856972 | 2012, 2013, 2014, 2015, 2016 | *Eidolon helvum*, *Epomophorus gambianus*, *Epomops franqueti*, *Epomops buettikoferi*, *Hypsignathus monstrosus*, *Epomophorus pusillus*, *Myonycteris angolensis*, *Nanonycteris veldkampii* | 657 |
| Ve-Golokwati | 6.996111, 0.437806 | 2012, 2013, 2014, 2015, 2016 | *Eidolon helvum*, *Epomophorus gambianus*, *Epomops franqueti*, *Epomops buettikoferi*, *Epomophorus pusillus*, *Nanonycteris veldkampii*, *Rousettus aegyptiacus* | 774 |
| Yendi | 9.461901, -0.008524 | 2015 | *Epomophorus gambianus* | 19 |
| Captive colony | 5.625303, -0.203029 | 2012, 2013, 2014, 2016, 2018, 2019 | *Eidolon helvum* | 1,639 |
| Bat samples (tested against EBOV, MARV, BDBV, RESTV, TAFV, SUDV, LLOV, MLAV, BOMV, and RAVV) | | | | |
| 37 Hospital, Accra | 5.5882, -0.18239 | 2019–2020 | *Eidolon helvum* | 496 |
| Kumasi | 6.70002, -1.62493 | 2019–2020 | *Eidolon helvum* | 433 |
| Akosombo | 6.171256, 0.064621 | 2019–2020 | *Eidolon helvum* | 386 |
| Captive colony | 5.625303, -0.203029 | 2019–2020 | *Eidolon helvum* | 775 |
| Human samples (tested against EBOV, MARV) | | | | |
| 37 Hospital, Accra | 5.5882, -0.18239 | 2014 | Humans (healthy volunteers, indirect or no bat contact) | 120 |
| 37 Hospital, Accra | 5.5882, -0.18239 | 2011 | Humans (febrile patients, unknown bat contact) | 79 |
| Urban Accra | 5.551379, -0.198623 | 2010 | Humans (indirect or no bat contact) | 152 |
| Akonkonti | 7.671122, -1.958847 | 2014 | Humans (cave exposure) | 61 |
| Amangoase | 7.671122, -1.958847 | 2014 | Humans (cave exposure) | 52 |

| | | | | |
|---|---|---|---|---|
| Bonya | 7.671122, -1.958847 | 2015 | Humans (cave exposure) | 26 |
| Buoyem | 7.671122, -1.958847 | 2011–2012, 2014 | Humans (cave exposure) | 139 |
| Forikrom | 7.592040, -1.857755 | 2011–2012 | Humans (cave exposure) | 152 |
| Kwamang | 6.975479, -1.272321 | 2011–2012 | Humans (cave exposure) | 203 |
| Tanoboase | 7.662612, -1.856934 | 2014 | Humans (hunting exposure) | 75 |
| Ve-Golokwati | 6.998066, 0.438414 | 2014 | Humans (cave exposure) | 106 |
| Volta | 7.2, 0.316667 | 2011 | Humans (hunting exposure) | 135 |

**Supplementary Table 2.** Smoothing and parametric terms included in GAMMs of lnMFI values for EBOV and MARV in wild *E. helvum*, *E. gambianus*, and *R. aegyptiacus* and their: effective degrees of freedom (edf), F values (*F*), and significance levels (*p*-value) for smoothing terms; and estimates, standard errors (*SE*), t statistics (*t* value), and significance levels (*p*-value) for parametric terms. The reference demographic status is adult male.

| **Antigen** | | | | | |
|---|---|---|---|---|---|
| EBOV | **Smoothing term** | | **edf** | ***F*** | ***p*-value** |
| | s(Time): *Eidolon helvum* | | 15.23 | 15.64 | <0.0001 |
| | s(Time): *Epomophorus gambianus* | | 19.24 | 24.69 | <0.0001 |
| | s(Time): *Rousettus aegyptiacus* | | 10.05 | 71.23 | <0.0001 |
| | **Parametric term** | **Estimate** | ***SE*** | ***t* value** | ***p*-value** |
| | Female (not pregnant) | -0.01 | 0.03 | -0.44 | 0.6587 |
| | Pregnant/lactating | 0.01 | 0.01 | 0.74 | 0.4613 |
| | Sexually immature | -0.02 | 0.01 | -2.51 | 0.0121 |
| | Body condition | -0.00 | 0.00 | -0.55 | 0.5806 |
| MARV | **Smoothing term** | | **edf** | ***F*** | ***p*-value** |
| | s(Time): *Eidolon helvum* | | 16.33 | 17.21 | <0.0001 |
| | s(Time): *Epomophorus gambianus* | | 19.25 | 35.59 | <0.0001 |
| | s(Time): *Rousettus aegyptiacus* | | 10.40 | 223.11 | <0.0001 |
| | **Parametric term** | **Estimate** | ***SE*** | ***t* value** | ***p*-value** |
| | Female (not pregnant) | -0.02 | 0.02 | -0.90 | 0.3670 |
| | Pregnant/lactating | -0.00 | 0.01 | -0.33 | 0.7450 |
| | Sexually immature | -0.03 | 0.01 | -5.98 | <0.0001 |
| | Body condition | -0.00 | 0.00 | -0.09 | 0.9330 |
